# Surgery versus external beam radiotherapy for hepatocellular carcinoma involving the inferior vena cava or right atrium: A systematic review and meta-analysis

**DOI:** 10.1101/2020.09.20.20198440

**Authors:** Han-Ah Lee, Sunmin Park, Yeon Seok Seo, Won Sup Yoon, In-Soo Shin, Chai Hong Rim

**Affiliations:** Department of Gastroenterology, Korea University Anam Hospital, Korea University Medical College, Seoul, Korea; Department of Radiation oncology, Korea University Ansan Hospital, Korea University Medical College, Gyeonggido, Korea; Graduation school of Education, Dongguk University, Seoul, Korea

**Keywords:** Hepatocellular carcinoma, Inferior vena cava, Meta-analysis, Radiotherapy, Surgery

## Abstract

**Purpose:** As the treatment efficacy of systemic therapy for patients with advanced-stage HCC is insufficient, locoregional therapies are performed in the clinical practice. We investigated the efficacy and safety of two most potent therapies, surgery and external beam radiotherapy (EBRT), in patients with HCC involving the IVC and/or right atrium (RA) through comparative meta-analysis.

**Method:** A systematic search of Pubmed, MEDLINE, EMBASE, and the Cochrane library was performed for entries up to July 2020. The primary endpoints were 1- and 2-year overall survival (OS) rates, while secondary endpoints were response rate, local control rate, and grade ≥3 toxicities.

**Results:** Eighteen studies with 22 cohorts were included, encompassing 755 patients. The pooled median OS and 1-and 2-year OS rates were 14.2 months, 55.6%, and 27.4%, respectively. The pooled median OS in the surgery and EBRT arms were 15.3 and 11.7 months, respectively. The pooled 1-year OS rate of the surgery arm was significantly higher than that of the EBRT arm (62.4%, 95% CI: 53.8–70.3 vs. 48.8%, 95% CI: 40.9–56.8; p=0.023). However, the 2-year OS rates were comparable (26.9%, 95% CI: 20.7–34.2 vs. 27.5%, 95% CI: 19.7–37.1; p=0.913). The pooled response and local control rates in the EBRT arm were 74.3% and 87.2%, respectively. In the surgical arm, the perioperative mortality and grade ≥3 complication rates were 0–7.6% and 3.9–67%, respectively. Grade ≥3 complications and radiation-induced liver disease were rarely observed in the EBRT arm.

**Conclusions:** Both surgery and EBRT are effective treatment options for patients with HCC involving IVC/RA invasion. Outcomes and safety should be further evaluated in well-controlled clinical trials.

## Introduction

HCC is the sixth most common cancer and fourth major cause of cancer-related deaths worldwide [1]. The prevalence of HCC varies among regions; approximately 75% of HCCs occur in East Asia, where the major risk factor is chronic hepatitis B infection [2]. In contrast to patients in other geographic areas, a considerable number of those in East Asia are diagnosed with advanced stage despite active HCC surveillance [3].

It is well known that patients with macrovascular invasion have poor prognoses, with median survival times of 6–8 months when untreated [4, 5]. Compared to the portal vein, HCC less frequently involves the IVC and right atrium (RA); under 4% of all HCC patients have such involvement [6-8] but experience worse outcomes with expected survival times of less than 3 months without treatment [9]. In addition, IVC tumor thrombi can flow into the heart and lungs, causing pulmonary embolisms and lung metastases [10-12].

Although most guidelines recommend systemic therapy as the standard treatment for advanced-stage HCC [13], such treatments provide minimal or modest clinical benefits [14, 15]. Furthermore, other local treatments such as surgical resection or external beam radiotherapy (EBRT) have produced comparable or better efficacies than sorafenib in several studies of patients presenting with major vascular invasion [16, 17]. Hence, the use of systemic therapy as the first-line treatment is not prevalent, and many clinicians have attempted to reduce the tumor burden with locoregional therapies in real-world practice [18].

A standardized treatment for HCC involving the IVC/RA has not been established owing to a limited number of studies to date. Following advances in surgical techniques, recent studies showed that surgical management of these patients produced better survival outcomes than non-surgical alternatives. However, since surgical approaches for this group of patients are invasive, complete resection is not always possible, and it is necessary to balance between effectiveness and avoiding complications [19-21]. Although transarterial chemoembolization (TACE) is the most common local treatment modality, its oncologic outcome is unsatisfactory given its median survival of 4.2–4.7 months [22-24]. Since EBRT is feasible for treating HCC lesions including those in major vessels, it has also been performed in patients with IVC/RA involvement and was found to be effective and safe in a recent meta-analysis, dispelling previous belief that EBRT can lead to pulmonary embolism or cardiac toxicity [17].

Although surgical resection and EBRT have been considered potent treatment options for patients with HCC who have IVC/RA invasion, adequate comparative studies have not been conducted. Therefore, we performed this comparative meta-analysis to evaluate the efficacies and feasibilities of both surgical approach and EBRT for HCC patients with IVC/RA invasion.

## Methods

### Database search and study selection

The present systematic review and meta-analysis was designed to answer *the following PICO question: “Are surgical approach and EBRT for HCC patients with IVC and/or RA invasion comparable options regarding oncologic efficacy and feasibility?*” Four databases (EMBASE, Medline, Pubmed, and the Cochrane library) were systematically searched for literature published up to July 8th, 2020. Publications prior to the year 2000 were excluded; no language restriction was applied. Conference abstracts were not considered owing to the lack of clinical data that compare between treatment modalities. Detailed search strategies, including the search terms used, are shown in Supplement 1.

Studies included in the present analyses met the following criteria: 1) clinical studies involving at least 10 patients with HCC who have IVC and/or RA invasion and were treated with either surgery or EBRT, and 2) survival outcomes were detailed. Multiple studies from the same institution were all included if patient overlap was either not present or negligible; if patient overlap was significant, the following criteria were used for selection, prioritized in the order listed: 1) studies involving HCC patients with IVC and/or RA invasion exclusively (i.e., not those in which such invasion were subgroups or prognosticators); 2) larger number of patients; and 3) more recently published.

After the initial search, studies with irrelevant formats (e.g., reviews, letters, conference abstracts, editorials, case reports, trial protocols, and lab studies) were excluded using machine filtering functions provided by EMBASE and PubMed. Duplicate studies were filtered out using the Rayan QCRI web-based program (https://rayyan.qcri.org/welcome). Initial searches across databases and machine filtering were performed by an experienced meta-analysis researcher (CHR). Next, abstract and full-text reviews were performed by 2 independent researchers to identify suitable studies. The studies that were finally included were verified by all authors.

### Data acquisition

Data were recorded on a pre-standardized coding sheet that included 1) general information (author names, affiliations, publication year, funding sources); 2) clinical data including age, etiology, Child-Pugh class, (CPC), alpha-fetoprotein level, accompanying PVT rate, extrahepatic metastasis rate, tumor size, radiotherapy (RT) method, complete resection rate, pre-, combined, or post-treatment; and 3) the primary endpoint of overall survival (OS) as well as secondary endpoints that included response rate after EBRT, first progression site, grade ≥3 complications, and perioperative mortality. In the absence of numeral data, OS rates were estimated using descriptive graphs. Two independent researchers acquired the data, and any disagreements were resolved with mutual discussion and a re-review of the literature.

### Quality assessment and statistical methods

Considering that most studies of patients with HCC are non-randomized clinical series, we used the Newcastle-Ottawa scale for quality assessment [25]. Studies with scores of 7 to 9 were considered high quality and those with scores of 4 to 6 were deemed medium quality. Considering that a vast majority of studies shared a similar design, no specific method to differentiate studies based on data synthesis (e.g., different weighting or subgroup analyses according to the studies’ qualities) was applied. This meta-analysis included studies at the outcome level. Although it targeted a rare type of HCC, there can still be significant distinctions in terms of clinical characteristics and treatment details; therefore, a random effects model was used for all pooled estimates.

All endpoints of interest were measured as rates (percentages), and pooled rates were calculated as the weighted averages of those rates. The exception was that grade ≥3 complications and perioperative mortality rates were analyzed qualitatively. Heterogeneity among studies was assessed using Cochran’s Q test [26] and I^2^ statistics [27]. I^2^ values of 25%, 50%, and 75% corresponded to low, moderate, and high heterogeneity, respectively. Publication bias was assessed by visual inspection of funnel plots and quantitative analyses using Egger’s test [28]. If significant asymmetry was observed and the 2-tailed p-value in Egger’s test was lower than 0.1, Duval and Tweedie’s trim and fill was performed for sensitivity analysis [29]. Subgroup comparisons were performed using mixed effects analyses and Cochran’s Q-test, based on analysis of variance. Subgroup analyses were performed to examine local treatment methods (surgery vs. EBRT) as well as known clinical prognosticators including CPC-A, accompanying PVT, and extrahepatic metastases [17, 30]. Borderline values between subgroups were set close to the overall median value. The p-values <0.05 were considered significant on subgroup analyses. Statistical analyses were performed by an experienced meta-analysis researcher (CHR) and supervised by a biostatistician specialized in meta-analyses (ISS). All statistical analyses were performed using Comprehensive Meta-Analysis version 3 (Biostat Inc., Englewood, NJ, USA).

## Results

### Study inclusion and general information

The initial database searches identified 1139 articles; however, initial machine filtering excluded 879 studies with irrelevant formats as well as 25 duplicates. Next, 235 studies underwent abstract screening; among them, 207 were excluded after investigation by 2 independent researchers. Full-text reviews of 28 articles were performed, and 18 studies of 22 cohorts involving 755 patients were ultimately included in the present study. The study selection process is shown in Figure 1.

**Figure 1.**
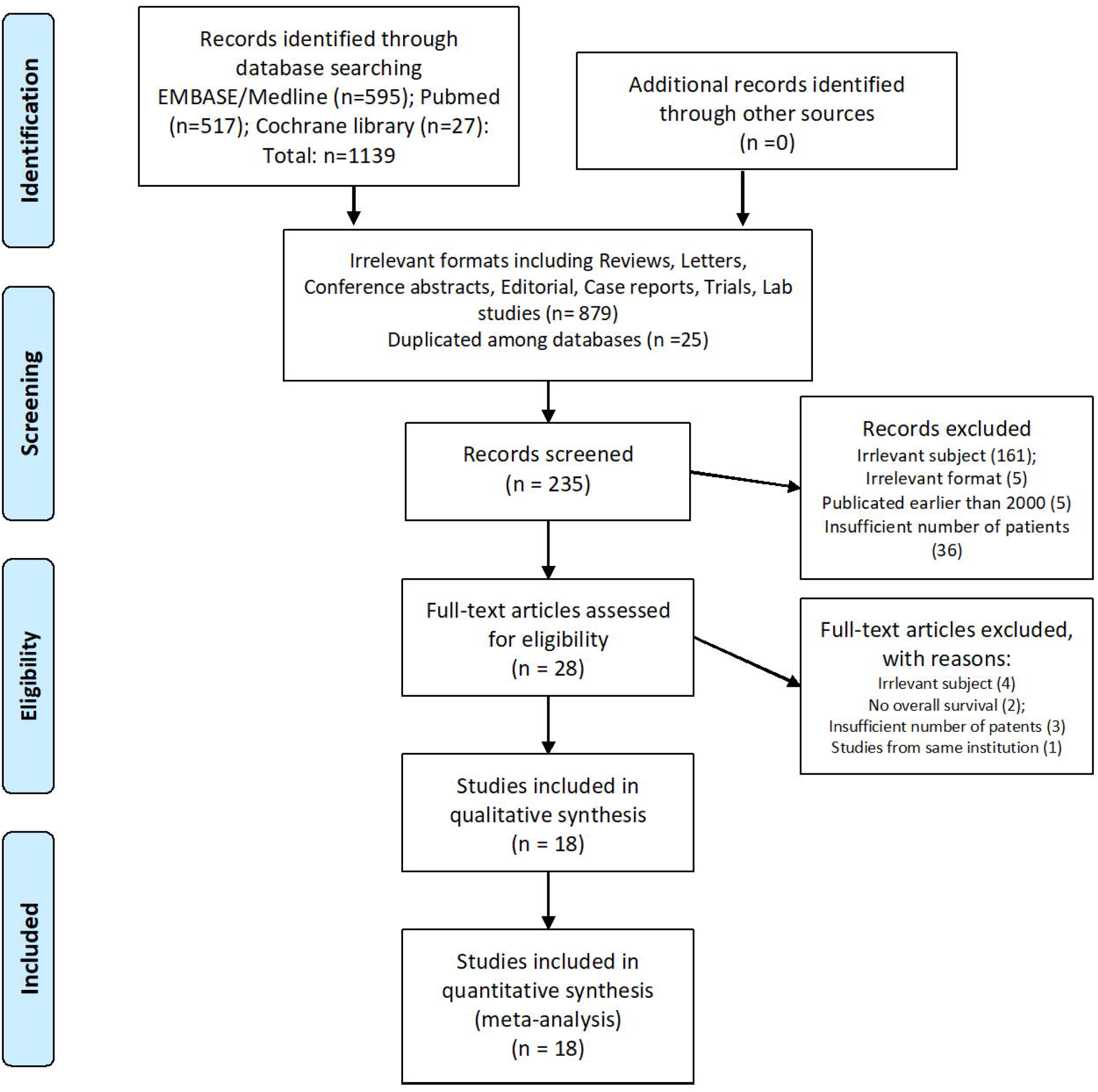
Study inclusion plot.

Among the 18 studies, 8 were single arm studies [19-22, 31-34] that were categorized into the surgery subgroup, another 8 were single arm studies [24, 35-41] categorized into the EBRT subgroup, and 2 were controlled studies [42, 43] comprising 2 cohorts that compared surgery and EBRT. Eight studies (44%) were from mainland China, 6 from Japan (33%), 2 from Korea (11%), and 1 from Taiwan (5.6%). A vast majority of patients in the studies performed in China, Korea, and Taiwan had HCC related to hepatitis B virus, while the proportions of patients with HCC related to hepatitis B and C viruses were comparable in studies from Japan. The rates of patients with CPC-A ranged from 72.9% to 100% (median: 100%) in the surgery arm and from 44.4% to 88.0% (median: 60.7%) in the EBRT arm. The median rates of PVT were 48.0% (range: 0%–100%) and 32.5% (range: 0%–100%) in the surgery and EBRT arms, respectively, while the corresponding rates of extrahepatic metastases (including lymph node metastasis) were 10.7% (range: 0–61.5%) and 30.8% (range: 0–94.6%), respectively. Data from the studies are shown in Table 1.

**Table 1.**
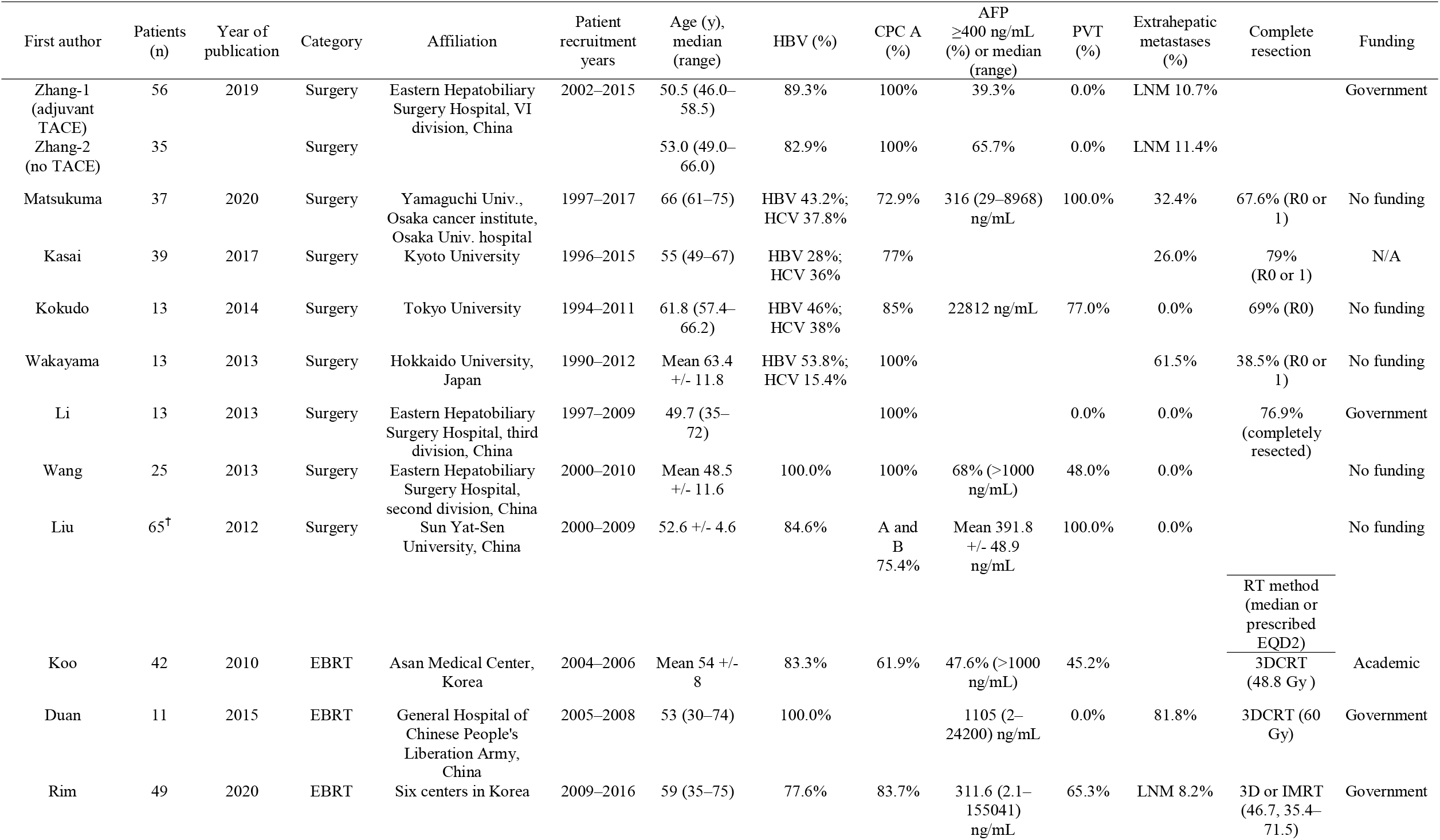
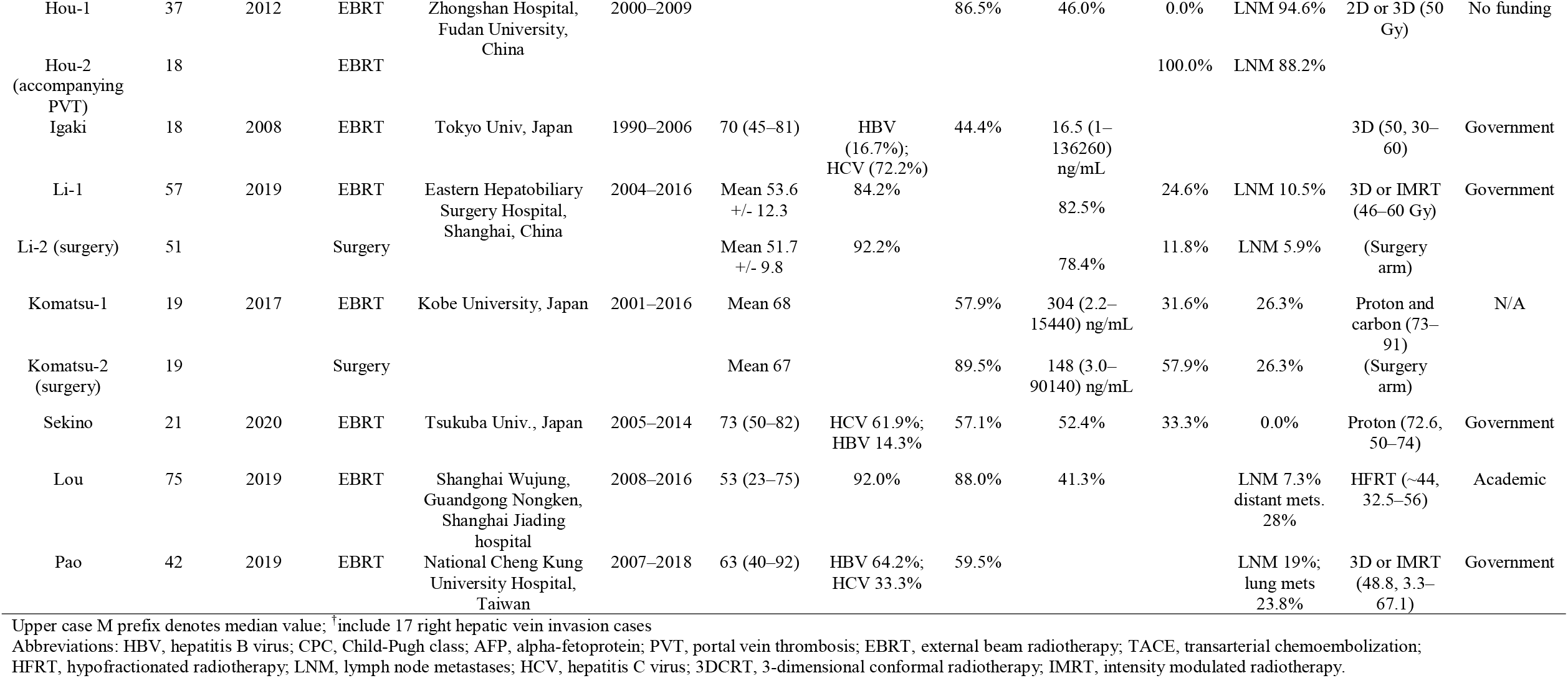
General data from the studies included in this meta-analysis.

### Quality analysis

All studies were deemed medium quality except for 2 controlled studies by Li et al. [42] and Komatsu et al. [43] that were high quality. All medium-quality studies lacked points for the selection of non-exposed cohorts and comparability between cohorts; detailed results are shown in Supplement 2. Funding information was available for 16 of the 18 studies. Eight studies were supported by the government, 2 by academic funding, and 6 received no funding. None of the studies reported conflicts of interest regarding commercial ties (Table 1).

### Clinical data and outcomes of interest

The median OS periods were 14.2 months (range: 5–21 months), 15.3 months (range: 13.0–19.0 months), and 11.7 months (range: 5.6–21.0 months) for all studies, the surgery arms, and EBRT arms, respectively. Among the 8 studies that reported prognostic factors related to OS, tumor multiplicity was reported in 4 studies, extrahepatic metastasis in 3, and CPC in 3. The most common sites of first progression were the liver (21.5–61.5%) and lung (13.5–42.9%). TACE and chemotherapy were the most common modalities applied pre- or post-surgery or radiotherapy. Li et al. (1 of the 2 controlled trials) reported that the median OS was similar in both the surgery and EBRT arms (14.5 vs. 12.8 months, p=0.466), while Komatsu et al. (the other controlled trial) demonstrated that the 1-year OS was lower in the surgical arm than in the EBRT arm with a non-significant trend (25% vs. 68%, p=0.106). Clinical data from the included studies are shown in Table 2.

**Table 2.**
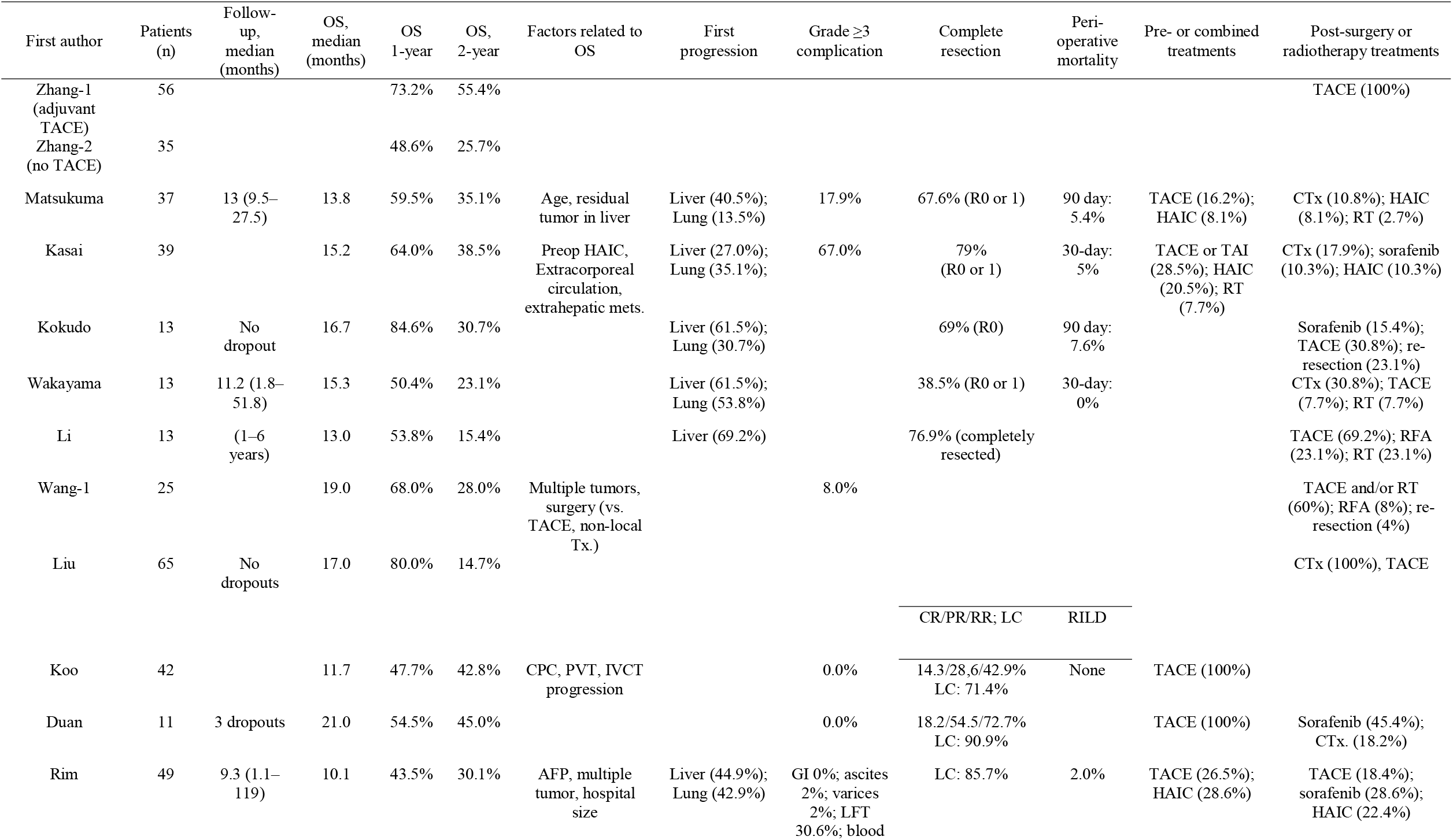
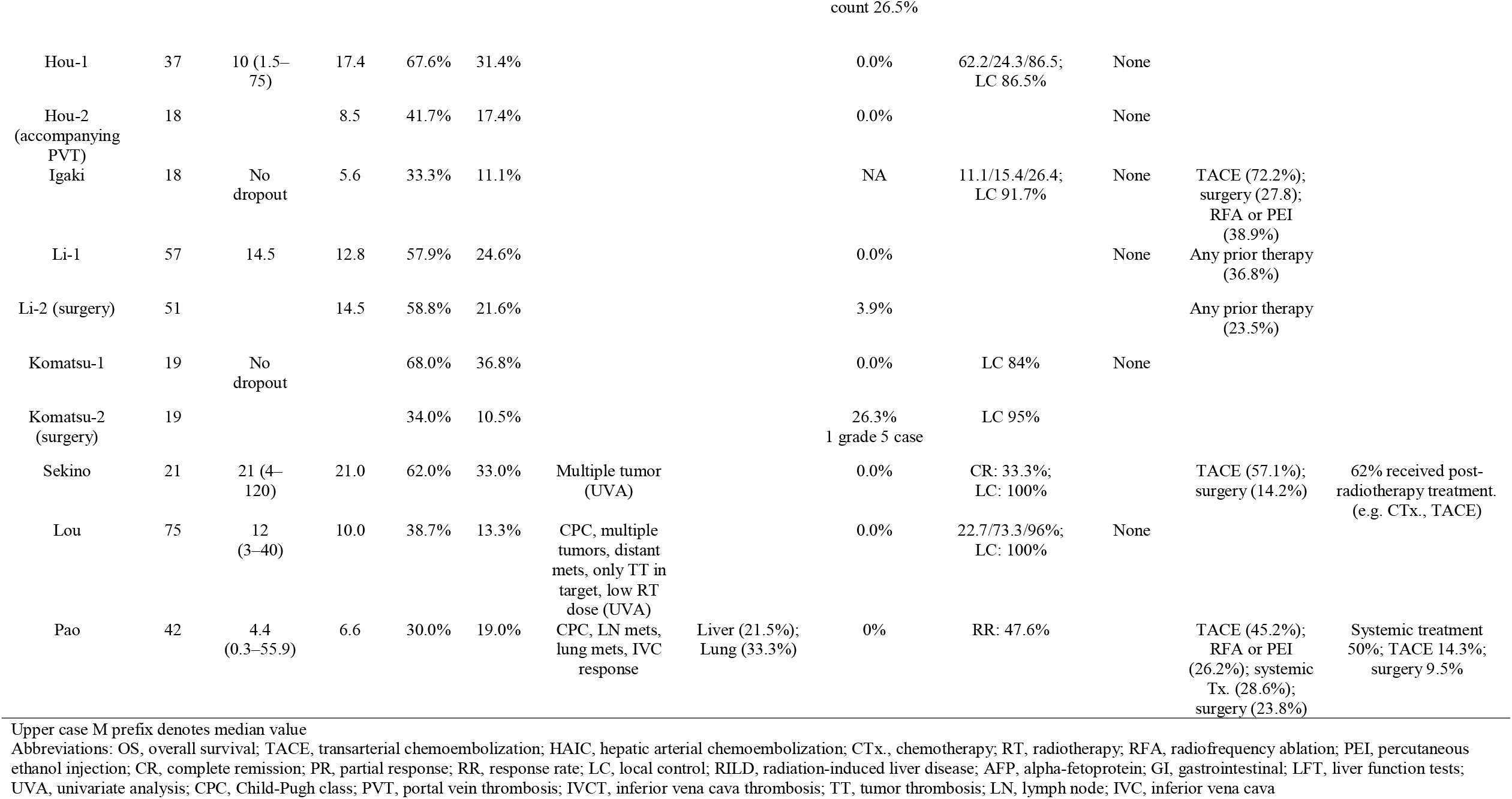
Clinical data of interest from the included studies.

The surgery arm had a higher pooled rate of patients with CPC-A than did the EBRT arm (90.1%, 95% CI: 80.2–95.4 vs. EBRT: 70.0%, 95% CI: 56.9–80.5; p=0.007) and had a lower rate of patients with extrahepatic metastases (14.5%, 95% CI: 7.6–25.8 vs. 34.5%, 95% CI: 16.5–58.3, p=0.067). The pooled 1- and 2-year OS rates for all studies were 55.6% (95% CI: 48.9–62.1) and 27.4% (95% CI: 22.3–33.1), respectively. The surgery arm had a higher 1-year OS rate (62.4%, 95% CI: 53.8–70.3) than did the EBRT arm (48.8%, 95% CI: 40.9–56.8) (p=0.023), although the 2-year OS rates were similar (26.9%, 95% CI: 20.7–34.2 vs. 27.5%, 95% CI: 19.7–37.1; p=0.913). On subgroup analyses, patients with lower extrahepatic metastasis rates (<20% vs. ≥20%) showed higher 1-year OS rates with borderline significance (62.7% vs. 50.6%, p=0.072). Pooled results are shown in Table 3 and depicted as forest plots in Figure 2.

**Table 3.** Pooled results of endpoints.

|  | No. of studies | No. of patients | Heterogeneity p | I<sup>2</sup> (%) | Heterogeneity | Pooled results (95% CI) | p-value (subgroup comparison) | Egger's p (trimmed value)* |
| --- | --- | --- | --- | --- | --- | --- | --- | --- |
| *Child Pugh class A (%)* |  |  |  |  |  |  |  |  |
| Surgery | 9 | 250 | 0.02 | 55.8% | High | 90.1% (80.2–95.4) | **0.007** |  |
| EBRT | 8 | 303 | <0.001 | 77.0% | Very high | 70.0% (56.9–80.5) |  |  |
| *Accompanying PVT (%)* |  |  |  |  |  |  |  |  |
| Surgery | 9 | 314 | <0.001 | 88.3% | Very high | 40.8% (14.3–74.1) | 0.815 |  |
| EBRT | 8 | 254 | <0.001 | 81.4% | Very high | 36.2% (20.0–56.3) |  |  |
| *Extrahepatic metastases (%)* |  |  |  |  |  |  |  |  |
| Surgery | 11 | 366 | <0.001 | 73.7% | High | 14.5% (7.6–25.8) | **0.067** |  |
| EBRT | 8 | 311 | <0.001 | 88.5% | Very high | 34.5% (16.5–58.3) |  |  |
| *1-year OS* |  |  |  |  |  |  |  |  |
| All studies | 22 | 755 | <0.001 | 66.4% | High | 55.6% (48.9–62.1) |  | 0.673 |
| Surgery | 11 | 389 | 0.009 | 57.7% | High | 62.4% (53.8–70.3) | **0.023** |  |
| EBRT | 11 | 366 | 0.014 | 55.0% | High | 48.8% (40.9–56.8) |  |  |
| PVT <50% | 11 | 367 | 0.361 | 8.7% | Low | 60.2% (54.7–65.4) | 0.801 |  |
| PVT ≥50% | 6 | 201 | <0.001 | 80.1% | High | 57.8% (40.0–73.8) |  |  |
| EHM <20% | 10 | 385 | 0.004 | 63.3% | High | 62.7% (53.8–70.9) | **0.072** |  |
| EHM ≥20% | 10 | 310 | 0.005 | 61.7% | High | 50.6% (40.8–60.3) |  |  |
| CPC A <80% | 7 | 218 | 0.014 | 62.5% | High | 51.8% (40.4–63.1) | 0.641 |  |
| CPC A ≥80% | 10 | 335 | 0.001 | 68.7% | High | 55.6% (44.9–65.7) |  |  |
| AFP <400 ng/mL<sup>†</sup> | 10 | 417 | <0.001 | 79.2% | Very high | 55.7% (44.2–66.6) | 0.617 |  |
| AFP ≥400 ng/mL | 7 | 213 | 0.469 | ~0.0% | Very low | 59.1% (52.2–65.6) |  |  |
| *2-year OS* |  |  |  |  |  |  |  |  |
| All studies | 22 | 755 | <0.001 | 59.3% | High | 27.4% (22.3–33.1) |  | 0.078 (32.3%) |
| Surgery | 11 | 366 | 0.001 | 67.4% | High | 26.9% (20.7–34.2) | 0.913 |  |
| EBRT | 11 | 389 | 0.031 | 49.6% | Moderate | 27.5% (19.7–37.1) |  |  |
| PVT <50% | 11 | 367 | 0.016 | 54.1% | High | 32.9% (25.8–40.9) | 0.121 |  |
| PVT ≥50% | 6 | 201 | 0.114 | 43.7% | Moderate | 23.5% (16.0–33.2) |  |  |
| EHM <20% | 10 | 385 | 0.002 | 66.1% | High | 27.8% (20.3–36.8) | 0.801 |  |
| EHM ≥20% | 10 | 310 | 0.034 | 50.2% | Moderate | 26.3% (19.4–34.6) |  |  |
| CPC A <80% | 7 | 218 | 0.152 | 36.3% | Moderate | 32.4% (24.8–41.1) | 0.366 |  |
| CPC A ≥80% | 10 | 335 | <0.001 | 69.7% | High | 26.5% (18.2–37.0) |  |  |
| AFP <400 ng/mL<sup>†</sup> | 10 | 417 | <0.001 | 78.2% | High | 27.5% (18.7–38.6) | 0.925 |  |
| AFP ≥400 ng/mL | 7 | 213 | 0.79 | ~0.0% | Very low | 27.0% (21.4–33.4) |  |  |

Abbreviations: CI, confidence interval; OS, overall survival; EBRT, external beam radiotherapy; PVT, portal vein thrombosis; EHM, extrahepatic metastases; CPC, Child-Pugh class; AFP, alpha-fetoprotein
\*Duval and Tweedie's trim and fill method for values with possible publication bias (Egger's p < 0.1)
<sup>†</sup>Median value of ≥400 ng/mL or more than 50% of patients had AFP levels of ≥400 ng/mL

**Figure 2.**
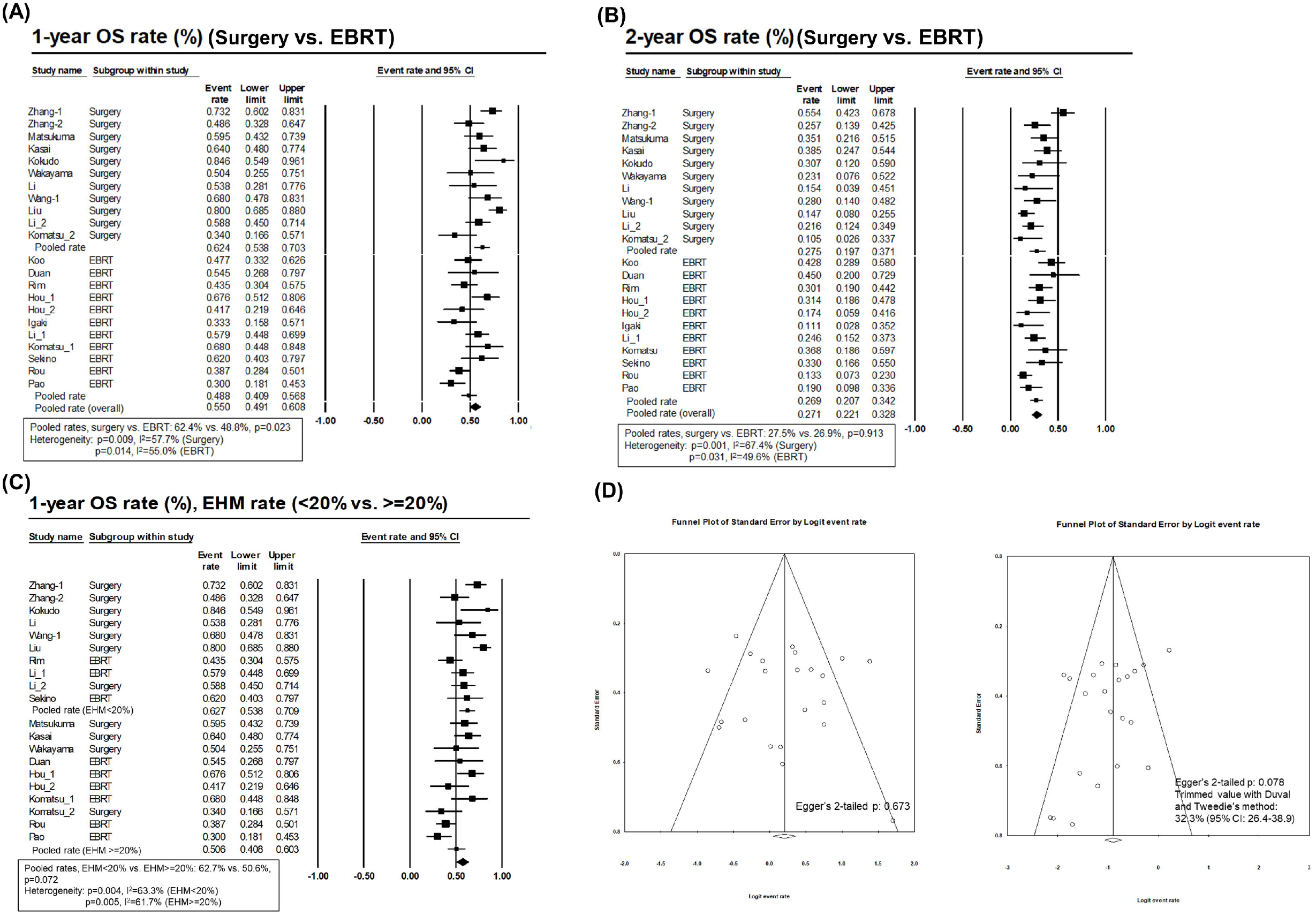
Forest plots of 1-year (A) and 2-year overall survival rates (B); forest plot of the 1-year overall survival rate according to extrahepatic metastasis rate (C); funnel plot of the 1-year (left) and 2-year overall survival rate on pooled analyses (D). Abbreviations: OS, overall survival; EBRT, external beam radiation therapy; EHM, extrahepatic metastasis

As for secondary endpoints, the most common sites of first progression were the liver (pooled rate: 43.2%, 95% CI: 31.0–56.2), followed by the lung (pooled rate: 34.0%, 95% CI: 24.6–44.9). The pooled rates of complete remission, response rate, and local control after EBRT were 27.9% (95% CI: 15.5–45.0), 74.3% (95% CI: 46.9–90.5), and 87.2% (95% CI: 78.2–92.8), respectively (Table 4). Four studies in the surgery arm reported either 30-day or 90-day perioperative mortality rates, which ranged from 0 to 7.6%. Grade ≥3 complication rates were reported in 5 surgery cohorts and ranged from 3.9% to 67%. Radiation-induced liver disease was very rare, having been reported in only 1 study (2% rate). Grade ≥3 complications in the EBRT arm were nonexistent (0%) in all cohorts except Rim et al.’s study [36], wherein the rates of gastrointestinal complications, ascites, and varices were 0%, 2%, and 2%, respectively. In that same study, the rates of transient elevation of liver function markers (e.g., albumin, bilirubin, and aspartate transaminase/alanine transaminase: 30.6%) and decrease in blood count (e.g., white blood cells, hemoglobin, and platelets: 26.5%) were relatively high (Table 2).

**Table 4.** Pooled results of selected secondary endpoints.

|  | No. of studies | No. of patients | Heterogeneity p | I<sup>2</sup> (%) | Heterogeneity | Pooled results (95% CI) |
| --- | --- | --- | --- | --- | --- | --- |
| *EBRT response rates (EBRT studies only)* |  |  |  |  |  |  |
| LC rate | 8 | 272 | 0.053 | 49.7% | Moderate | 87.2% (78.2–92.8) |
| CR rate | 5 | 186 | 0.002 | 76.3% | Very high | 27.9% (15.5–45.0) |
| RR rate | 5 | 207 | <0.001 | 89.8% | Very high | 74.3% (46.9–90.5) |
| *First progression site (all available studies)* |  |  |  |  |  |  |
| Liver | 7 | 206 | 0.007 | 65.8% | High | 43.2% (31.0–56.2) |
| Lung | 6 | 193 | 0.074 | 50.2% | Moderate | 34.0% (24.6–44.9) |

Abbreviations: CI, confidence interval; EBRT, external beam radiotherapy; LC, local control; CR, complete remission; RR, response rate

### Heterogeneity and publication bias analyses

Heterogeneity among cohorts was moderate-to-high in most of the pooled analyses except on subgroup analyses of 1- and 2-year OS rates in patients with high alpha-fetoprotein and of the 1-year OS rate in the patients with low PVT rates. Potential publication bias was not evident in the pooled analysis of the 1-year OS rate, but was noted for that of the 2-year OS rate; the 2-tailed p-value in Egger’s test was 0.078, and the trimmed value using Duval and Tweedie’s method was increased from the original value (pooled 2-year OS rate: 27.4% to 32.3%) (Table 3). Funnel plots are shown in Figure 2.

## Discussion

Patients with HCC that includes IVC/RA invasion have a poor prognosis and risk life-threatening complications. Although pioneering researchers have applied active local treatments to improve clinical outcomes, previous data were not sufficient to derive optimal treatment strategies. In this comparative meta-analysis, we evaluated the therapeutic effect and safety of the 2 most promising treatment modalities, surgical resection and EBRT, for patients with HCC involving the IVC/RA.

Our study had several clinical strengths and implications. Our pooled median OS was 14.2 months, which is much longer than that of previous series [22-24, 44, 45]. Chun et al. [44] showed that the median OS of patients with IVC/RA invasion was 4 and 2 months in the actively treated and supportive care groups, respectively. In the active treatment group, only 18.7% of patients underwent surgery or EBRT, whereas approximately half of the patients underwent 5-fluorouracil-based chemotherapy or TACE. When compared to the median OS in studies of patients treated with TACE (4.2–4.7 months) [22-24], the treatment efficacy of surgery and EBRT appeared to be superior to that of TACE alone. In the same vein, Wang et al. [22] compared the survival benefits of surgery and TACE in a matched cohort comparison; the median survival times were 19 vs. 4.5 months (p<0.001). Koo et al. [24]. also demonstrated that combined TACE and EBRT yielded higher survival than the control group that underwent TACE alone (median survival: 11.7 vs. 4.7 months, p<0.01).

The median survival of patients in the surgery arm in our study was 15.3 months, which was longer than 11.7 months in the EBRT arm; this suggests that surgery might produce a greater survival benefit than EBRT for these patients. However, caution is needed when interpreting this result because there were significantly more patients with CPC-A (90.1% vs. 70.0%, p=0.007) while extrahepatic metastases tended to be less common (14.5% vs. 34.5%, p=0.067) in the surgery arm. Impaired liver function and the presence of extrahepatic metastasis are known predictors of poor prognosis in patients with advanced-stage HCC [46]. Therefore, to reduce the effects of selection bias and confounding factors on survival, a well-designed propensity score-matched study is required. Indeed, in the study by Li et al. that compared the treatment effects of surgery and RT in patients without extrahepatic metastasis and with similar characteristics between groups, no significant difference in survival was observed (median survivals with surgery vs. EBRT: 14.5 vs. 12.8 months, p=0.466) [21]. A study by Komatsu et al. also showed comparable survival outcomes (p=0.106) in the surgery and EBRT groups following matched-pair analysis that adjusted for 12 clinical factors [43].

In contrast to the significantly higher 1-year OS rate in the surgery arm than in the EBRT arm (62.4% vs. 48.8%, p=0.023), the 2-year OS rates in the 2 groups were comparable (26.9% vs. 27.5%, p=0.913). Although most surgeries aim to remove all tumor lesions, advanced-stage HCCs may not be completely resectable and can remain macroscopic tumor or microscopic metastases. Even after curative resection, early recurrence (which characteristically occurs within 2 years after treatment) is frequent in patients with advanced-stage HCC [47, 48], and this may contribute to considerable differences in mortality between 1 and 2 years among those who undergo surgical resection [49, 50]. Indeed, 1-year recurrence-free survival in studies where surgery was applied ranged from 26% to 44% [19, 20, 31, 32].

The pooled response and local control rates following EBRT (the secondary outcomes of our study), were 74.3% and nearly 90%, respectively, demonstrating potent local control with EBRT. With the introduction of 3-dimensional conformal radiation therapy, it has become possible to apply therapeutic doses of 45–60 Gy to the tumor while sparing the normal liver. Therefore, the treatment efficacy for liver malignancies has markedly increased [51-53]. EBRT can be safely applied for tumors that involve major vessels, with a tolerance dose of over 100 Gy, whereas other modalities may be contraindicated owing to possible hemorrhage [54]. Objective response or local control of IVC/RA after EBRT was shown to be significantly related to survival [24, 41]. Therefore, EBRT could be an effective treatment modality, possibly enhancing the oncologic outcomes of these patients. Additionally, the pooled response and local control rates of HCCs that invaded the IVC/RA were higher than those of HCCs with PVTs in previous studies [55]. This phenomenon could be explained by the difference between the portal and hepatic veins, as the components of the PVT (tumor and blood cells that block the narrow-diameter vessels) are less affected by EBRT than are the isolated tumor thrombi in the IVCs [37, 56].

Although the surgery arm demonstrated higher median and 1-year survival rates, the grade ≥3 complication rates in this arm were higher than those in the EBRT arm (3.9–67% vs. 0–4%); moreover, post-operative mortality rates of 0–7.6% were observed in the former. For radical resection of HCCs involving the IVC/RA, concerns exist regarding the burden on the circulatory system that arises owing to invasive procedures, such as total hepatic vascular exclusion or extracorporeal circulation. Furthermore, pulmonary embolisms and pleural effusions, which are closely associated with circulatory dysfunction, commonly develop [19, 20, 22]. There are also concerns that EBRT for treating IVC/RA thromboses might cause systemic complications, such as pulmonary embolism or cardiac failure. However, most studies in the EBRT arm showed an absence or a very low rate of grade ≥3 complications (including radiation-induced liver disease). This may be due to successfully limiting the bystander radiation dose to adjacent major organs (such as the lung and heart) to levels lower than the standardized dose constraints [13, 57]. In addition, duodenal toxicity, which is a common and serious complication when treating HCC with EBRT, is less a concern because of the longer distance between the IVC/RA thrombosis and duodenum [54]. Given these safety issues, surgery could be considered for patients with favorable clinical characteristics, whereas EBRT could be a less invasive alternative option that has potent treatment efficacy.

Lastly, the most common sites of first progression after local control using EBRT were the liver (43.2%) and lung (34.0%), as expected. In the multi-institutional study by Rim et al. [36], which is one of the largest EBRT series, the median progression-free survival after EBRT was only 4 months in patients with HCC exhibiting IVC/RA invasion. After surgical resection, the median recurrence-free survival only reached 5.2–5.3 months [19, 20, 31, 32]. When considering the short time before the disease progressed, local control may not be sufficient to prevent further progression in substantial patients with advanced-stage HCC, therefore, there is a distinct need for preventing further HCC progression after local treatment. Studies evaluating the effect of systemic therapy after local control in advanced-stage HCC are scarce. Rim et al. reported that patients who had concurrent or additional systemic treatment had longer median overall survival than those without such treatment (12.2 vs. 8.4 months, p=0.054); however, the trend was not significant [17]. Kasai et al. evaluated the effect of adjuvant sorafenib therapy after surgical resection in patients with HCC involving the IVC/RA and showed comparable OS and recurrence-free survival between patients with and without adjuvant therapy [20].

Recently, the immune-related effects of RT have gained attention in the era of immunotherapy because RT may have pro-immunogenic effects on immune responses [58]. Preclinical data demonstrated that RT could improve the response of cancer cells to immune checkpoint inhibitors by enhancing immunogenic antigen presentation, stimulating proinflammatory cytokines, and increasing PD-L1 expression [59]. The results of several ongoing clinical trials that are evaluating the effect of EBRT alone or in combination with systemic therapies could provide valuable perspectives to clinicians.

We are also aware of several limitations that remained unresolved. Meta-analyses of non-randomized studies are controversial because the pooled effects can be influenced by different study designs and patient characteristics [60]. Regarding our present study, detailed methods of surgery or radiotherapy varied among institutions, and there would have been differences in the application of additional treatments aside from the main local modalities. Although a large randomized trial might provide robust evidence with minimal biases, the field of oncology does not always provide the best evidence, and clinical decisions are commonly based on small or observational studies [61]. HCC involving the IVC/RA is a very rare condition with a poor prognosis; therefore, a meta-analysis of observational studies might be one of the few available methods to assess treatment efficacy and feasibility.

In conclusion, both surgery and EBRT are effective treatment options for patients with HCC exhibiting IVC/RA invasion. Surgical resection appears to provide a longer survival benefit than EBRT; however, its higher rate of complications should also be considered. EBRT could be safely used in patients with HCC involving the IVC/RA as a feasible and effective palliative treatment option. Further studies comparing the efficacy and safety of surgical resection and EBRT in well-controlled patient groups are warranted.

## Data Availability

all data analyzed in this study is published in literature

## Declaration

### Ethical considerations

Ethical approval was not required because this study retrieved and synthesized data from papers that are already published.

## Author contributions

**Han Ah Lee**, writing-original draft, writing-review & editing; **Sunmin Park**, data curation, writing-review & editing; **Yeon Seok Seo**, supervision**; Won Sup Yoon**, supervision**; In-Soo Shin**, methodology; **Chai Hong Rim**, conceptualization, writing-original draft, writing-review & editing, formal analysis, data curation

## Abbreviations

CI: confidence interval
CPC: Child-Pugh class
EBRT: external beam radiotherapy
IVC: inferior vena cava
HCC: hepatocellular carcinoma
OS: overall survival
PVT: portal vein thrombosis
RA: right atrium
TACE: transarterial chemoembolization

